# Weight Loss Dynamics and Health Burden Changes with Tirzepatide versus Semaglutide

**DOI:** 10.64898/2025.11.30.25341294

**Authors:** AJ Venkatakrishnan, Karthik Murugadoss, Venky Soundararajan

## Abstract

**Background:** Glucagon-like peptide-1 receptor agonists (GLP-1RAs) are widely used for obesity and type 2 diabetes, yet substantial variability in weight-loss response remains poorly understood.

**Methods:** We conducted a retrospective cohort study using de-identified electronic health records and performed 1:1 propensity matching on age, sex, type 2 diabetes status, baseline BMI and weight, index year, and follow-up duration. The matched cohorts included 10,339 tirzepatide-treated and 10,339 semaglutide-treated patients. Patients were categorized by maximum weight loss over two years into five response groups. Adverse events were identified through AI-enabled curation of clinical notes. Weight-loss trajectories, demographic patterns, adverse-event profiles, and pre- to post-treatment disease-prevalence changes were compared across drugs.

**Results:** Patients treated with tirzepatide lost more weight than those treated with semaglutide (mean reduction, 14.7% vs. 10.8%; p<0.001). High-response rates (≥15% weight loss in year 1) were nearly doubled with tirzepatide (42.6% vs. 21.6%; p<0.001), accompanied by faster monthly weight-loss velocity (2.54% vs. 2.18%). AI-enabled curation showed that tirzepatide was associated with lower prevalence of gastrointestinal and systemic adverse events. For both tirzepatide and semaglutide, women were more represented among high responders than the minimal weight-loss group (<5% weight loss) and White patients were more represented among high responders, whereas Black and Hispanic patients were more represented among the minimal weight loss group.

**Conclusions:** In this large, propensity-matched real-world cohort, tirzepatide was associated with greater and faster weight loss than semaglutide. Marked demographic disparities highlight the need for precision approaches to obesity treatment.

## Introduction

Incretin-based therapies have transformed obesity treatment by producing clinically meaningful weight loss, yet real-world responses span a wide range^1^. Clinical trials and observational cohorts often emphasize average results, but the magnitude of interindividual variability suggests distinct biological subgroups that remain insufficiently defined^2,3^. The characteristics of exceptional responders, the clinical profiles of non-responders, the pattern of adverse events, and the real-world duration of therapy are not fully understood^3,4^. Electronic health records permit large-scale, longitudinal characterization of weight trajectories and comorbid conditions, although skepticism regarding data quality necessitates careful attention to confounding, population heterogeneity, and ascertainment bias^5–7^. Artificial intelligence applied to unstructured notes can augment structured data and improve capture of clinical events^5,8,9^. In this study, by leveraging propensity-score matching and AI-based curation of clinical documentation, we characterize response heterogeneity, identify physiologic signatures across response groups, and examine differential patterns of adverse events and disease burden changes across tirzepatide and semaglutide.

## Methods

### Data Source

This study analyzed de-identified EHR data from academic medical centers in the United States via the nference nSights Analytics Platform. Prior to analysis, all data underwent expert determination de-identification satisfying HIPAA Privacy Rule requirements (45 CFR §164.514(b)(1)), employing a multi-layered transformation approach for both structured data (cryptographic hashing of identifiers, date-shifting, geographic truncation) and unstructured clinical text (ensemble deep learning and rule-based methods with >99% recall for personally identifiable information detection)^10,11^. nference established secure data environments within each participating center, housing these de-identified patient data governed by expert determination. These de-identified data environments were specifically designed to enable data access and analysis without requiring Institutional Review Board oversight, approval, or exemption confirmation. Accordingly, informed consent and IRB review were not required for this study.

### Cohort Selection

From a longitudinal database of 23 million patients, we identified 640,794 with at least one GLP-1RA order. Requiring ≥3 orders spanning ≥1 month yielded 278,471 patients; restricting to semaglutide or tirzepatide prescriptions yielded 215,004 patients. We constructed drug-exclusive cohorts comprising patients who received only semaglutide brands (Ozempic, Wegovy, Rybelsus) or only tirzepatide brands (Mounjaro, Zepbound); patients could switch between brands of the same drug but not between drugs. We excluded patients with bariatric surgery history and required weight measurements during baseline (≤90 days pre-treatment), the first year post-treatment (year 1), and the second year post-treatment (year 2), yielding 34,143 semaglutide-treated and 16,034 tirzepatide-treated patients (**Figure S1**).

### Weight Trajectories

Patients in both semaglutide and tirzepatide cohorts were stratified into five response groups (**Figure 1**): high response (>15% weight loss during the entire study period), moderate response (5-15% during the entire study period), late response (<5% during year 1, >5% during year 2), weight regain (>5% during year 1, ≥5% regain during year 2), and minimal weight loss (<5% weight loss during the entire study period). The index date was the date of the first GLP-1RA prescription Baseline weight was the measurement closest to the index date within 90 days pre-treatment. Weight changes were calculated relative to baseline and aggregated into 30-day windows (±15 days). Population-level trajectories were generated by computing patient-level mean percentage changes within each window, then smoothed using Savitzky-Golay filtering (window=5, polynomial order=3). Uncertainty was quantified as standard error of the mean (SEM=σ/√n); shaded regions represent ±1 SEM.

**Figure 1.**
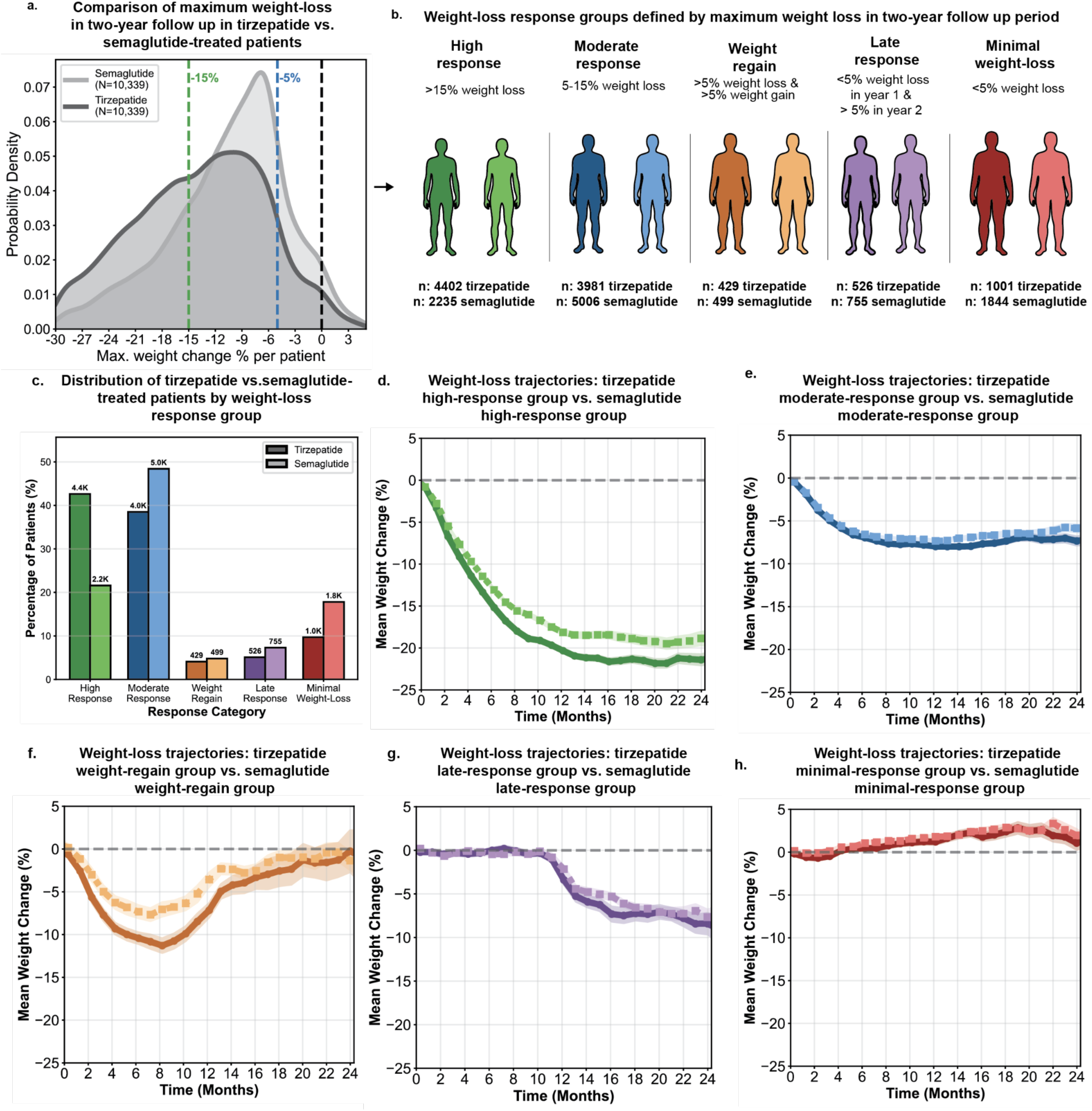
Comparative Weight-Loss Response Profiles and Longitudinal Trajectories in Propensity-Matched Tirzepatide- and Semaglutide-Treated Patients. Panel A shows the distribution of maximum weight loss over a two-year follow-up among tirzepatide-treated and semaglutide-treated patients after 1:1 propensity matching (N=10,339 per cohort), illustrating a right-shifted distribution for tirzepatide consistent with greater overall weight reduction. Panel B defines the five weight-loss response categories based on maximum percentage weight change: high response (>15% loss), moderate response (5–15% loss), weight-regain group (>5% loss in year 1 followed by >5% regain in year 2), late response (<5% loss in year 1 but >5% cumulative loss in year 2), and minimal weight loss (<5% loss). The corresponding counts of tirzepatide- and semaglutide-treated patients in each group are shown. Panel C compares the distribution of patients across response categories, demonstrating a higher proportion of high responders with tirzepatide and a higher proportion of minimal responders with semaglutide. Panels D through H present mean longitudinal weight-change trajectories over 24 months for tirzepatide versus semaglutide within each response category (high, moderate, weight-regain, late, and minimal response). Across categories, tirzepatide consistently produces steeper early weight loss and greater sustained reduction, whereas semaglutide shows attenuated initial decline and earlier plateauing. Error bands denote 95% confidence intervals.

### Propensity Score Matching

To compare treatment responses between tirzepatide and semaglutide while controlling for baseline differences, we performed propensity score matching for the overall tirzepatide and semaglutide populations as well as separately within each of the five weight-loss outcome categories (high response, moderate response, weight regain, late response, and minimal weight-loss groups). Propensity scores were estimated using multivariable logistic regression with treatment assignment (tirzepatide vs. semaglutide) as the dependent variable and the following covariates: age at index date, sex, type 2 diabetes mellitus status (defined as ≥3 diagnoses within 5 years prior to index), baseline body mass index (most recent measurement within 90 days before index), baseline weight (most recent measurement within 90 days before index), year of treatment initiation, and follow-up duration in days. Nearest-neighbor matching without replacement was performed using a 1:1 matching ratio and a caliper width of 0.1 standard deviations of the logit of the propensity score. Patients with index dates before January 1, 2023 were excluded to ensure comparable follow-up periods and account for differential market availability of the medications. Balance diagnostics were assessed by comparing standardized mean differences for all matching covariates between matched cohorts within each response category, with values <0.1 considered indicative of adequate balance. We characterized brand name distributions within matched cohorts (**Tables S1-S2**) among the tirzepatide (Mounjaro, Zepbound) and semaglutide (Ozempic, Wegovy, Rybelsus) branded formulations.

### Statistical Analysis

Summary statistics were generated for semaglutide and tirzepatide cohorts across all response groups (**Tables S3-S8**). Demographics included age at first prescription, gender, and race/ethnicity. Race and ethnicity data were derived from patient self-report documented in the electronic health record. We categorized patients as White (non-Hispanic), Black/African American (non-Hispanic), Hispanic (any race), or Other/Unknown, with the latter category combining all other racial groups and missing data due to small sample sizes that precluded separate statistical analysis^12^. Baseline clinical characteristics included Type 2 diabetes mellitus (T2DM), body mass index (BMI), and weight. BMI and weight were defined as the nearest recorded values within 90 days prior to the index date. T2DM status was defined by at least three diabetes diagnostic codes recorded in the 5 years prior to the index date. We characterized weight measurement distributions across a 10-year pre-treatment period and the first two years post-treatment. GLP-1RA prescription intervals were calculated as the time between first and last prescription for each patient (**Figure S2**), and prescription frequency was defined as the total number of prescriptions received per patient. Follow-up duration was computed as days from index date to last clinical encounter for that patient. Baseline and post-treatment monthly clinical note counts were quantified. Statistical comparisons between propensity-matched tirzepatide and semaglutide cohorts were performed using two-proportion z-tests for binary outcomes and chi-square tests or Fisher’s exact tests (for cell counts <5) for categorical outcomes.

### AI-enabled Augmented Curation

We employed a multi-stage information extraction (IE) pipeline using fine-tuned Bidirectional Encoder Representations from Transformers (BERT)^13^ models to extract clinical phenotypes from unstructured clinical notes. The pipeline first applies Named Entity Recognition (NER) to identify clinical entities, then uses sequential qualifier models to assess clinical context by determining subject (patient vs. other), temporality (current, past, or hypothetical), and certainty (confirmed, negated, or suspected). This contextualization framework disambiguates confirmed current patient symptoms from negated findings, suspected conditions, or prior diagnoses. This IE approach has been validated in prior studies^8^ with F1 scores of 0.93 and 0.95 for conditions and medications, respectively.

### Post-treatment Adverse Event Prevalence Analysis

We performed a comparative analysis of adverse event prevalence between propensity-matched tirzepatide and semaglutide cohorts within each weight-loss response category. We analyzed 16 commonly reported adverse events identified from FDA prescribing information for both medications, including gastrointestinal, systemic, and metabolic events. Adverse event prevalence was determined exclusively from unstructured clinical notes during the two-year post-treatment period using the validated information extraction pipeline described above. The pipeline identified adverse event mentions from all available clinical documentation, restricting output to confirmed, current patient-attributed events while excluding negated, suspected, or historical mentions. For each adverse event within each response group, we compared the prevalence in tirzepatide versus semaglutide cohorts using chi-square tests for adequate cell counts (≥5 expected events per cell) or Fisher’s exact test for smaller counts. We performed 16 independent tests within the high response, moderate response and minimal weight-loss categories separately and reported unadjusted p-values at p<0.05.

### Disease Prevalence Change Analysis

We performed a comprehensive analysis of pre- to post-treatment disease prevalence changes across 1,161 diseases extracted through AI curation from unstructured clinical text. To ensure robust statistical power, we prioritized diseases with ≥250 tirzepatide-treated patients in the pre-treatment period. Using the same validated information extraction pipeline described above, we identified disease mentions from all available clinical documentation in two-year pre-treatment and post-treatment time windows, restricting output to confirmed, current patient-attributed conditions. For each disease within each response group, we calculated prevalence ratios (pre-treatment prevalence divided by post-treatment prevalence) representing fold-reductions in disease burden for the propensity-matched tirzepatide and semaglutide cohorts. Statistical significance of pre- to post-treatment changes was assessed using chi-square tests. Given the exploratory nature of this analysis, we applied Bonferroni correction for all disease-category combinations tested (1,161 diseases × 5 response categories = 5,805 tests), with significance determined at corrected p<0.05. For patients with pre-treatment asthma and depression diagnoses, we additionally compared disease-specific changes in the pre-treatment versus post-treatment prevalence of structured depression or asthma related-medication prescriptions using the chi-square test as described above.

## Results

### Comparison of magnitude and rate of weight reduction in patients treated with tirzepatide versus semaglutide

In propensity matched cohorts of 10,339 patients across two years of follow-up, tirzepatide was associated with markedly greater maximal weight reduction than semaglutide (**Figure 1, Table S3-S5**). The mean of maximal two-year weight reduction was 14.7% (16.0 kg) for patients treated with tirzepatide compared to 10.8% (11.6 kg) for patients treated with semaglutide (p<0.001, two-proportion z-test, **Figure 1a**). Further, the proportion achieving their highest recorded weight loss of ≥5% at any time point was 90% with tirzepatide and 82% with semaglutide (p<0.001). Differences widened at higher thresholds: 68% versus 47% achieved ≥10% loss (p<0.001), and 43% versus 22% achieved ≥15% loss (p<0.001), respectively (**Figure 1a-c**). Maximal reductions of ≥20% were observed in 26% of tirzepatide-treated patients compared with 12% of semaglutide-treated patients (p<0.001), and ≥25% reductions were observed in 14% versus 5% (p<0.001) (**Figure 1a-c**). We compared the rate of weight loss between tirzepatide and semaglutide across all the weight-loss groups (**Figure 1d-h**). During the first six months after treatment initiation, tirzepatide was associated with more rapid weight reduction than semaglutide in the high-response group (**Figure 1d**). The mean monthly rate of weight loss was 2.54% for tirzepatide and 2.18% for semaglutide (p<0.001) in the high-response group. Taken together, these findings indicate that compared to semaglutide tirzepatide produced overall higher weight loss and among high responders faster weight loss.

### Demographic patterns in weight-loss response in patients treated with tirzepatide versus semaglutide

We evaluated the weight-loss response distributions by race/ethnicity and sex for both tirzepatide and semaglutide (**Table S3**). White individuals were more represented in the high-response group than the minimal response group for both tirzepatide (91.2% vs. 79.8%, p<0.001, two-proportion z-test) and semaglutide (91.2% vs. 79.1%, p<0.001). On the contrary, Black patients were more represented in the minimal weight-loss group than the high-response group for both tirzepatide (10.9% vs. 6.0%, p<0.001) and semaglutide (10.6% vs. 5.8%, p<0.001), with a similar pattern observed among Hispanic patients for tirzepatide (3.3% vs. 1.6%, p<0.001) and semaglutide (4.1% vs. 1.3%, p<0.001). Evaluating response distribution by sex, females were more represented in the high-response group than the minimal response group for both tirzepatide (74.9% vs. 59.2%, p<0.001) and semaglutide (80.3% vs. 58.5%, p<0.001).

### Comparison of prevalence of adverse events in patients treated with tirzepatide versus semaglutide

We compared the prevalence of 16 commonly reported adverse events between propensity-matched cohorts. In the high response group, the post-treatment prevalence of multiple gastrointestinal phenotypes was significantly lower among patients treated with tirzepatide compared to semaglutide, including nausea (27.2% versus 31.4%, p=0.002), vomiting (12.8% versus 17.1%, p<0.001), constipation (16.9% versus 20.2%, p=0.005), diarrhea (16.9% versus 19.2%, p=0.04), and dyspepsia (0.7% versus 1.6%, p=0.006). Additionally, tirzepatide-treated patients in the high response group had lower prevalence of fatigue (26.1% versus 29.7%, p=0.007) and headache (23.5% versus 28.4%, p<0.001). In the moderate response groups (N=2,962; **Figure 2b, Table S10**), tirzepatide was also associated with a lower post-treatment prevalence of nausea (16.4% versus 19.4%, p=0.003), vomiting (7.8% versus 9.6%, p=0.02), and dizziness (8.9% versus 11.0%, p=0.007).

**Figure 2.**
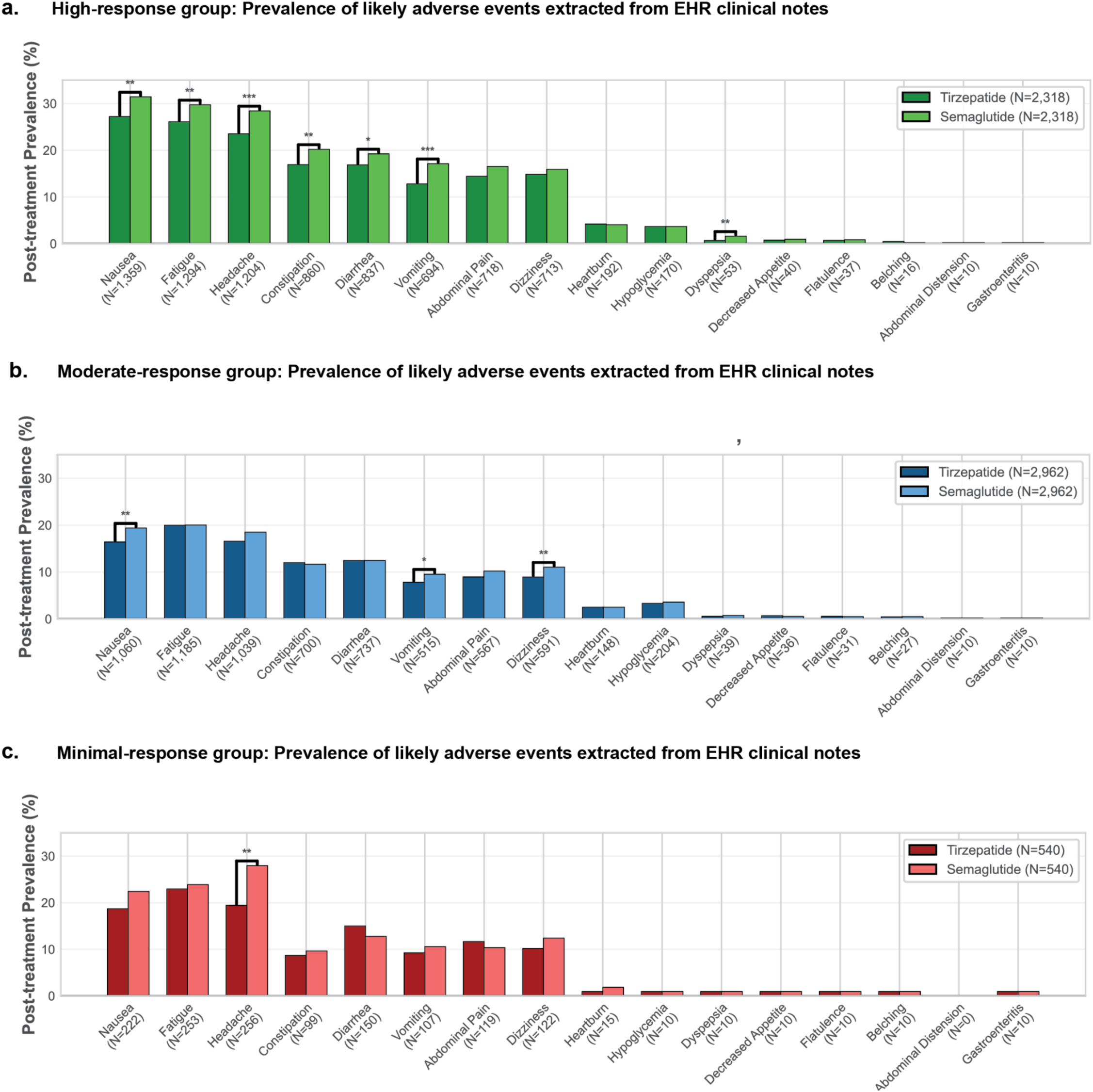
Prevalence of Adverse Events Extracted from Unstructured Clinical Text in Tirzepatide- and Semaglutide-Treated Patients Across Weight-Loss Response Groups. Panel A shows the post-treatment prevalence of selected adverse events among patients in the high-response group (N=2,318 per treatment cohort), as identified from unstructured clinical notes. Panel B presents corresponding adverse-event prevalence estimates for the moderate-response group (N=2,962 per cohort). Panel C shows prevalence estimates for the minimal-weight-loss group (N=540 per cohort). For each adverse event, bars represent the proportion of patients with at least one post-treatment mention; asterisks indicate the significance of between-treatment differences.

## Discussion

In this large, propensity-matched real-world comparative analysis of tirzepatide and semaglutide, tirzepatide was associated with greater weight loss overall and faster weight loss trajectories, with nearly twice the proportion of patients achieving ≥15% reductions. These findings extend observations from randomized trials^3,14^ and other real-world studies^15,16^ in highlighting diversity in weight-loss outcomes. Our findings underscore the profound heterogeneity of real-world outcomes: even after controlling for baseline characteristics, patients exhibited a wide distribution ranging from minimal change to >25% loss. Such dispersion illustrates the limitations of population-level efficacy metrics and the need for individualized treatment frameworks.

Demographics disparities in sex and race/ethnicity associated with weight-loss response were mirrored across both tirzepatide and semaglutide, suggesting that the observed distribution is not specific to a single agent. Although the underlying contributors to these differences cannot be determined from observational data, potential factors may include variation in baseline clinical characteristics, access to care, treatment persistence, dose escalation, social determinants of health, and unmeasured biological heterogeneity. The consistency of these response distributions across two distinct GLP-1RA therapies underscores the importance of further investigation into the multidimensional factors that shape weight-loss outcomes in diverse populations.

Differences between tirzepatide and semaglutide extended beyond weight loss. Patients treated with tirzepatide demonstrated consistently lower post-treatment prevalence of gastrointestinal and systemic adverse events across response strata, despite achieving higher magnitude and faster weight reductions. This dissociation suggests that tolerability may depend not solely on total weight loss or dose intensity, but on mechanistic distinctions between dual incretin agonism and GLP-1–only therapy. Improved tolerability among high responders may facilitate sustained adherence, contributing to the broader response distribution observed for tirzepatide.

AI-enabled extraction from unstructured clinical documentation enabled a high-resolution evaluation of changes in disease burden across 1,161 conditions. Both tirzepatide and semaglutide were associated with significant reductions in multiple disease categories, with 65 and 72 conditions reaching significance in high and moderate responders, respectively (Figure **S3a–b; Tables S12–S13**). Tirzepatide demonstrated superior improvements across several upper-respiratory conditions—including rhinorrhea, nasal congestion, and earache—where fold-reductions were consistently larger than those observed with semaglutide. Notably, both agents produced significant reductions in depression and asthma prevalence, accompanied by decreased antidepressant and asthma medication usage (**Tables S14–S17**), suggesting clinically meaningful improvement in symptomatic disease burden. These findings raise the possibility that GLP-1RA–mediated weight loss may confer secondary health benefits that extend beyond cardiometabolic parameters and warrant mechanistic investigation.

This study has limitations inherent to retrospective observational analyses, including potential residual confounding, incomplete adherence information, and reliance on provider documentation for disease and adverse event ascertainment. Nevertheless, the integration of two-year outcomes, rigorous propensity matching, millions of clinical notes analyzed through validated AI models, and one of the largest GLP-1RA-treated cohorts studied to date provides a robust characterization of real-world treatment heterogeneity.

In summary, tirzepatide was associated with greater and faster weight loss, lower adverse event prevalence, and larger reductions in multiple disease burdens compared with semaglutide, while both agents produced meaningful improvements across metabolic, respiratory, and neuropsychiatric domains. The wide spectrum of outcomes and demographic gradients observed emphasize the urgent need for precision approaches to obesity treatment and highlight opportunities for next-generation incretin therapeutics informed by real-world response phenotypes.

## Data Availability

This study involves the analysis of de-identified Electronic Health Record (EHR) data via the nference nSights Federated Clinical Analytics Platform (nSights). Data shown and reported in this manuscript were extracted from this environment using an established protocol for data extraction, aimed at preserving patient privacy. The data has been de-identified pursuant to an expert determination in accordance with the HIPAA Privacy Rule. Any data beyond what is reported in the manuscript, including but not limited to the raw EHR data, cannot be shared or released due to the parameters of the expert determination to maintain the data de-identification. Contact the corresponding author for additional details regarding nSights.

## Supplementary Material

**Figure S1:**
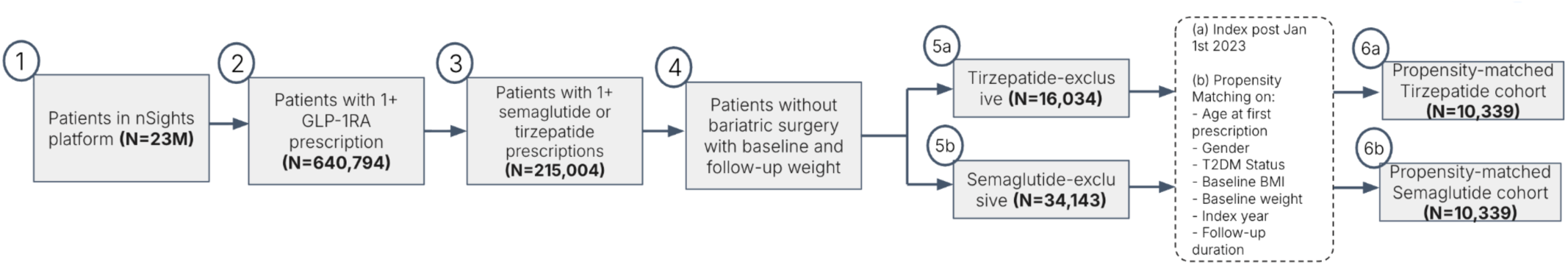
Cohort Funnel based on inclusion and exclusion criteria.

**Figure S2:**
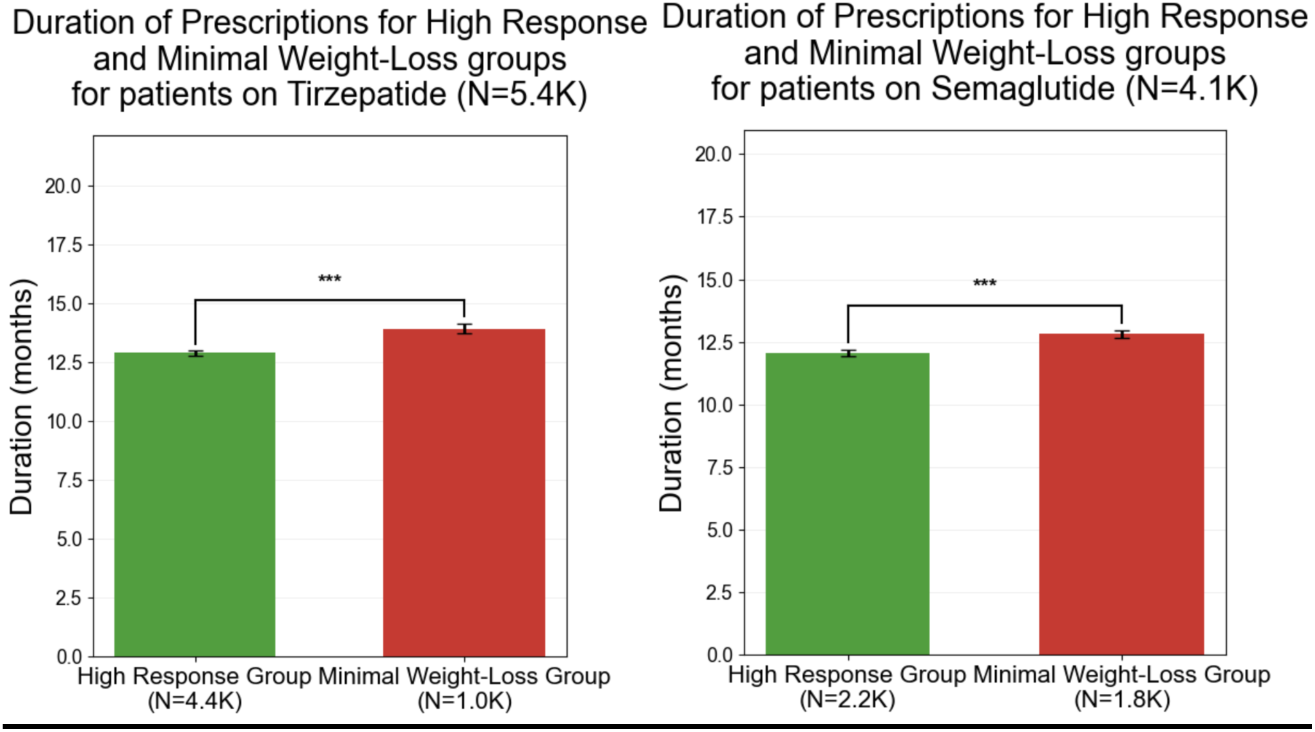
Distribution of duration of prescriptions (time from first to last prescription) in patients treated with (a) tirzepatide and (b) semaglutide.

**Figure S3.**
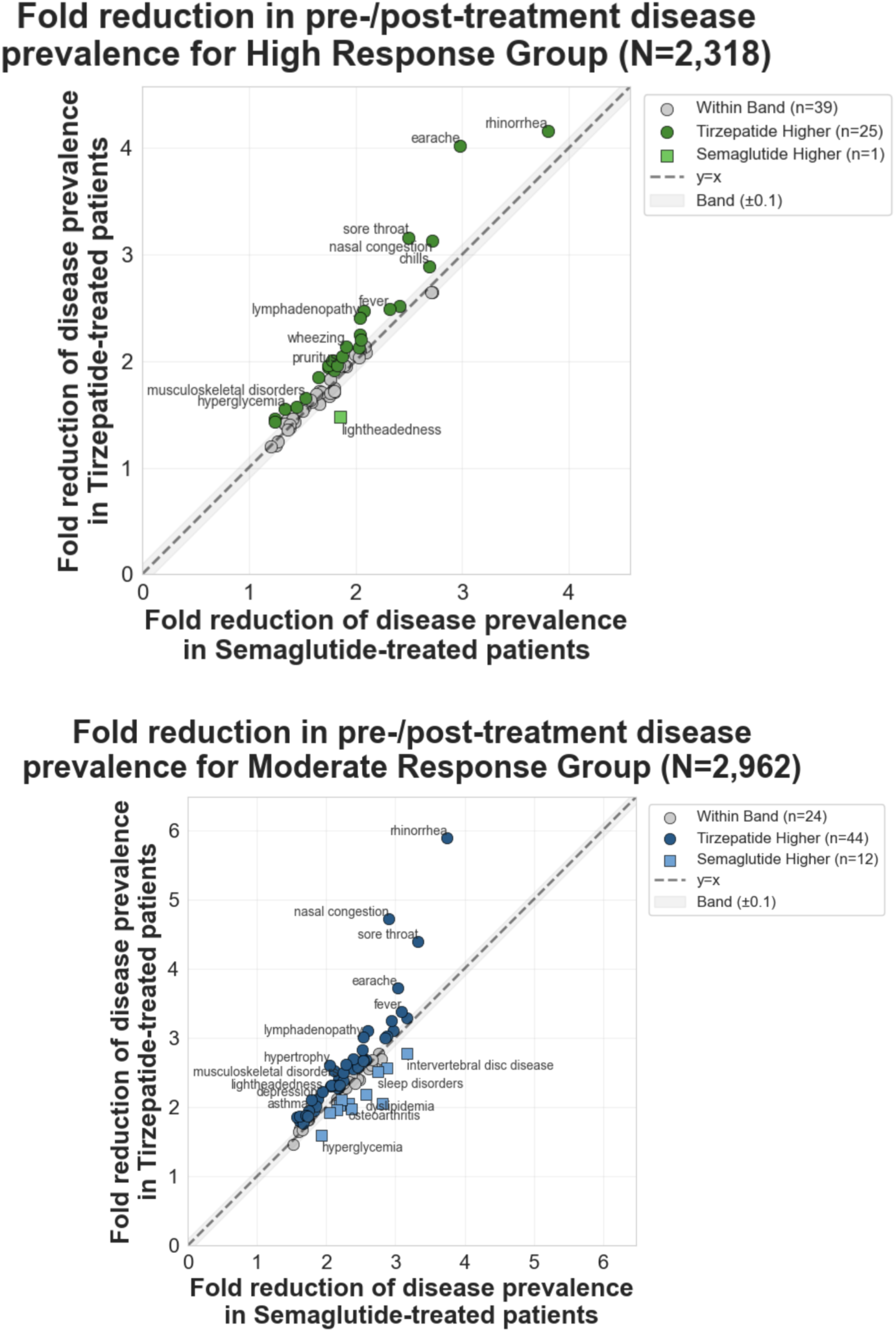
Comparative disease prevalence reductions following tirzepatide versus semaglutide treatment. Panel A (High Response Group, N=2,318): Pre- to post-treatment disease prevalence fold-reductions comparing tirzepatide (y-axis) versus semaglutide (x-axis). Each point represents one disease; points above the diagonal indicate greater reduction with tirzepatide. Green circles denote tirzepatide showing ≥0.1 greater fold-reduction, gray circles denote equivalent reductions within ±0.1 band, and green squares denote semaglutide showing ≥0.1 greater fold-reduction. Panel B (Moderate Response Group, N=2,962): Similar analysis showing amplified differential effects in moderate responders.

**Table S1.**
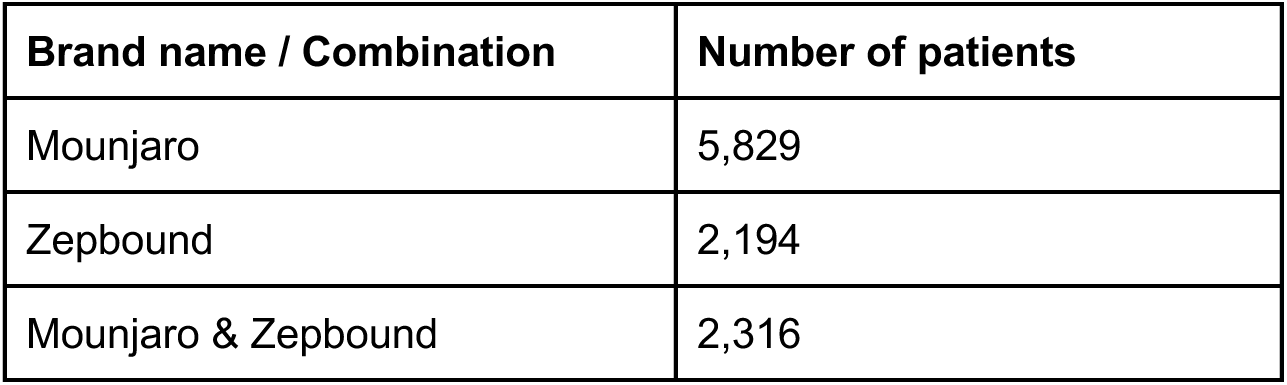
Brand name distribution among propensity-matched tirzepatide-treated patients (N=10,339)

**Table S2.** Brand name distribution among propensity-matched semaglutide-treated patients (N=10,339)

| **Brand name / Combination** | **Number of patients** |
| --- | --- |
| Ozempic | 5,851 |
| Wegovy | 2,862 |
| Ozempic & Wegovy | 850 |
| Rybelsus | 429 |
| Ozempic & Rybelsus | 294 |
| Wegovy & Rybelsus | 35 |
| Ozempic & Wegovy & Rybelsus | 18 |

**Table S3.** Demographic characteristics of propensity-matched tirzepatide vs semaglutide cohorts across the high response group, moderate response group, late response group, weight regain group and the minimal weight-loss group.

| DRUG<br>(Active Compound) | BRAND<br>(Marketed Formulation) | Weight-Loss Outcome Category | Number of Patients N (%) | Male (N) | Female (N) | Male Percentage (%) | Gender Association $-\log_{10}(P)$ | Age in years, Mean (SD) | Age in years, Median [IQR] | Race: White N (%) | Race: Black/African American N (%) | Race: Hispanic N (%) | Race: Other/Unknown N (%) |
| --- | --- | --- | --- | --- | --- | --- | --- | --- | --- | --- | --- | --- | --- |
| Semaglutide | (Ozempic/Wegovy/<br>Rybelsus) | High Response Group | 2235 (21.6%) | 439 | 1796 | 19.64 | 180.47 | 51.80 (14.93) | 51.47 [40.66, 63.37] | 2039 (91.2%) | 130 (5.8%) | 29 (1.3%) | 108 (4.8%) |
| Semaglutide | (Ozempic/Wegovy/<br>Rybelsus) | Moderate Response Group | 5006 (48.4%) | 1925 | 3081 | 38.45 | 59.28 | 56.08 (14.36) | 57.25 [46.66, 66.83] | 4371 (87.3%) | 382 (7.6%) | 125 (2.5%) | 252 (5.0%) |
| Semaglutide | (Ozempic/Wegovy/<br>Rybelsus) | Weight Regain Group | 499 (4.8%) | 177 | 322 | 35.47 | 10.07 | 52.64 (15.18) | 53.39 [40.60, 63.75] | 427 (85.6%) | 48 (9.6%) | 8 (1.6%) | 23 (4.6%) |
| Semaglutide | (Ozempic/Wegovy/<br>Rybelsus) | Late Response Group | 755 (7.3%) | 281 | 474 | 37.22 | 11.67 | 53.40 (13.99) | 54.34 [43.65, 64.07] | 634 (84.0%) | 60 (7.9%) | 24 (3.2%) | 50 (6.6%) |
| Semaglutide | (Ozempic/Wegovy/<br>Rybelsus) | Minimal Weight-Loss Group | 1844 (17.8%) | 766 | 1078 | 41.54 | 12.43 | 52.64 (14.52) | 53.69 [41.83, 64.28] | 1458 (79.1%) | 196 (10.6%) | 76 (4.1%) | 137 (7.4%) |
| Tirzepatide | (Mounjaro/Zepbound) | High Response Group | 4402 (42.6%) | 1108 | 3294 | 25.17 | 237.34 | 52.14 (13.26) | 52.38 [42.69, 62.28] | 4014 (91.2%) | 265 (6.0%) | 70 (1.6%) | 162 (3.7%) |
| Tirzepatide | (Mounjaro/Zepbound) | Moderate Response Group | 3981 (38.5%) | 1666 | 2315 | 41.85 | 24.09 | 55.09 (13.38) | 55.87 [45.46, 64.93] | 3480 (87.4%) | 272 (6.8%) | 81 (2.0%) | 227 (5.7%) |
| Tirzepatide | (Mounjaro/Zepbound) | Weight Regain Group | 429 (4.1%) | 142 | 287 | 33.1 | 11.59 | 52.25 (13.88) | 52.93 [42.47, 62.24] | 378 (88.1%) | 22 (5.1%) | 16 (3.7%) | 18 (4.2%) |
| Tirzepatide | (Mounjaro/Zepbound) | Late Response Group | 526 (5.1%) | 194 | 332 | 36.88 | 8.75 | 55.28 (12.61) | 55.98 [47.03, 64.28] | 457 (86.9%) | 35 (6.7%) | 14 (2.7%) | 24 (4.6%) |
| Tirzepatide | (Mounjaro/Zepbound) | Minimal Weight-Loss Group | 1001 (9.7%) | 408 | 593 | 40.76 | 8.3 | 52.56 (13.20) | 53.03 [43.04, 62.23] | 799 (79.8%) | 109 (10.9%) | 33 (3.3%) | 66 (6.6%) |

**Table S4.** Baseline clinical characteristics and data availability in propensity-matched tirzepatide versus semaglutide cohorts: Type 2 diabetes, BMI, weight distributions, weight measurement availability, and monthly clinical documentation across the high response group, moderate response group, late response group, weight regain group and the minimal weight-loss group.

| DRUG<br>(Active Compound) | BRAND<br>(Marketed Formulation) | Weight-Loss Outcome Category | With T2DM (N) | Without T2DM (N) | T2DM Percentage (%) | T2DM Association – log<sub>10</sub>(P) | Baseline BMI in kg/m<sup>2</sup>, Mean (SD) | Baseline BMI in kg/m<sup>2</sup>, Median [IQR] | Baseline Weight in kg, Mean (SD) | Baseline Weight in kg, Median [IQR] | Pre-GLP1RA Weight Measurements, Mean (SD) | Pre-GLP1RA Weight Measurements, Median [IQR] | Baseline monthly documents per patient, Mean (SD) | Baseline monthly documents per patient, Median [IQR] |
| --- | --- | --- | --- | --- | --- | --- | --- | --- | --- | --- | --- | --- | --- | --- |
| Semaglutide | (Ozempic/Wegovy/Rybelsus) | High Response Group | 477 | 1758 | 21.34 | 160.96 | 37.01 (6.09) | 36.39 [32.42, 41.71] | 104.54 (22.19) | 101.64 [88.47, 117.71] | 33.91 (32.68) | 25 [11, 45] | 6.3 (14.0) | 3 [2, 6] |
| Semaglutide | (Ozempic/Wegovy/Rybelsus) | Moderate Response Group | 1713 | 3293 | 34.22 | 109.74 | 36.90 (6.62) | 36.40 [32.15, 41.38] | 107.52 (23.37) | 104.64 [90.53, 122.20] | 34.61 (42.25) | 25 [11, 46] | 6.5 (14.0) | 3 [2, 6] |
| Semaglutide | (Ozempic/Wegovy/Rybelsus) | Weight Regain Group | 150 | 349 | 30.06 | 18.29 | 36.02 (6.84) | 35.25 [31.14, 41.47] | 104.70 (24.69) | 100.83 [86.87, 119.88] | 36.63 (37.39) | 26 [12, 49] | 8.3 (19.6) | 3 [2, 7] |
| Semaglutide | (Ozempic/Wegovy/Rybelsus) | Late Response Group | 241 | 514 | 31.92 | 22.53 | 37.91 (7.33) | 37.05 [32.59, 42.07] | 110.44 (24.44) | 108.76 [92.61, 125.46] | 29.36 (27.97) | 22 [11, 39] | 6.6 (13.5) | 3 [1, 7] |
| Semaglutide | (Ozempic/Wegovy/Rybelsus) | Minimal Weight-Loss Group | 568 | 1276 | 30.8 | 60.35 | 37.63 (7.63) | 36.80 [32.39, 42.13] | 110.60 (26.10) | 107.32 [92.36, 126.10] | 29.31 (30.23) | 21 [9, 39] | 7.0 (15.3) | 3 [1, 7] |
| Tirzepatide | (Mounjaro/Zepbound) | High Response Group | 1072 | 3330 | 24.35 | 253.14 | 37.53 (6.42) | 36.94 [32.78, 41.95] | 107.39 (22.75) | 104.07 [90.54, 121.36] | 31.15 (30.92) | 23 [11, 42] | 6.0 (13.4) | 3 [1, 6] |
| Tirzepatide | (Mounjaro/Zepbound) | Moderate Response Group | 1362 | 2619 | 34.21 | 87.58 | 36.88 (6.69) | 36.34 [32.06, 41.27] | 108.52 (24.09) | 106.16 [91.24, 123.01] | 30.72 (29.47) | 23 [11, 42] | 6.1 (13.7) | 3 [1, 6] |
| Tirzepatide | (Mounjaro/Zepbound) | Weight Regain Group | 122 | 307 | 28.44 | 18.38 | 35.59 (6.34) | 34.82 [31.10, 39.06] | 101.79 (22.23) | 97.82 [86.46, 115.09] | 30.70 (29.00) | 24 [10, 39] | 6.6 (12.9) | 3 [1, 7] |
| Tirzepatide | (Mounjaro/Zepbound) | Late Response Group | 177 | 349 | 33.65 | 13.19 | 38.03 (7.38) | 37.59 [33.07, 41.92] | 111.70 (24.67) | 107.97 [93.07, 126.87] | 28.84 (29.48) | 21 [11, 37] | 6.8 (14.6) | 3 [1, 6] |
| Tirzepatide | (Mounjaro/Zepbound) | Minimal Weight-Loss Group | 272 | 729 | 27.17 | 46.57 | 38.23 (9.18) | 37.48 [31.51, 43.64] | 112.69 (29.68) | 109.51 [91.49, 131.97] | 25.82 (23.57) | 20 [9, 35] | 6.5 (13.2) | 3 [1, 6] |

**Table S5.**
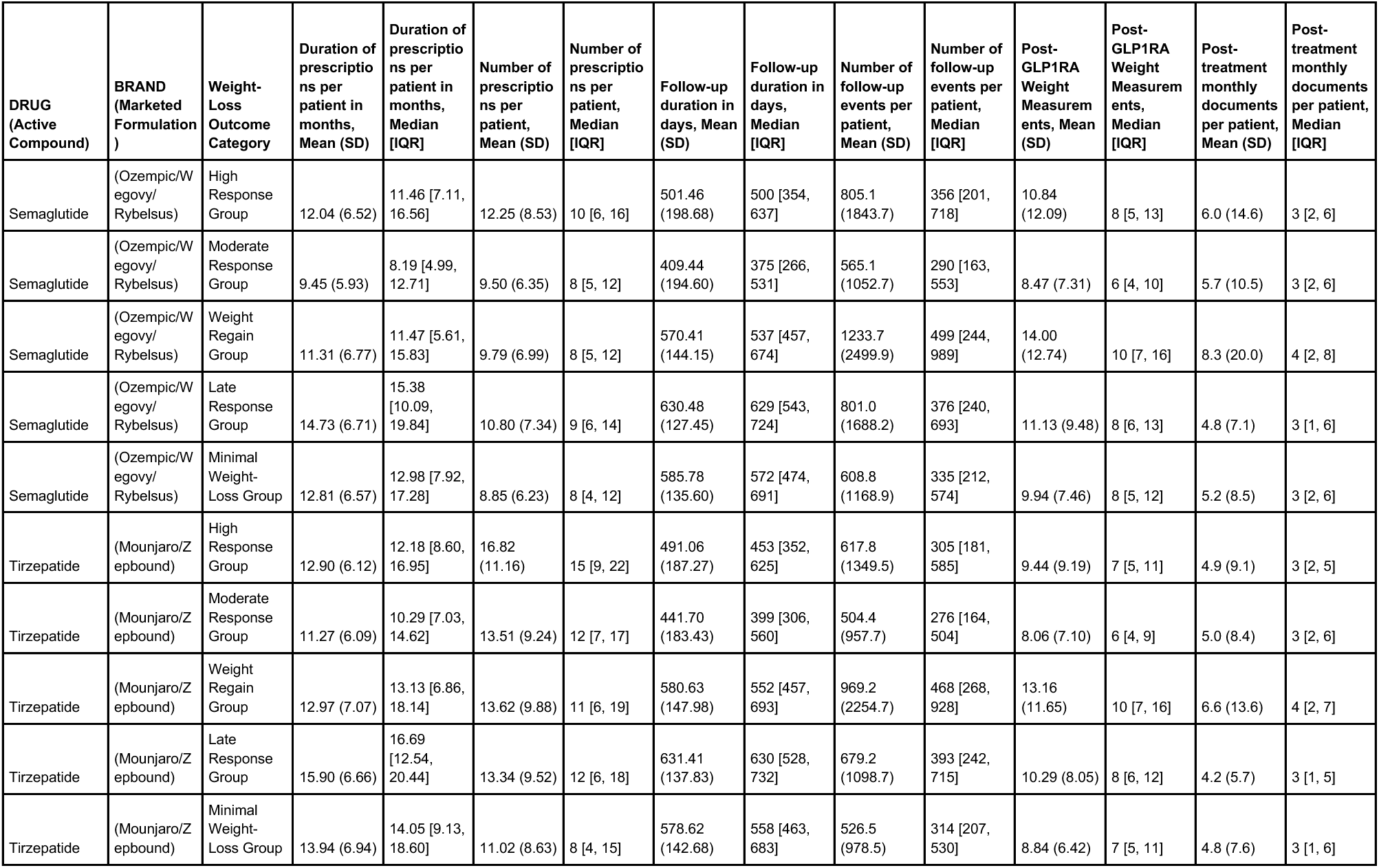
Post-treatment characteristics in propensity-matched tirzepatide versus semaglutide cohorts: prescription duration and frequency, follow-up duration, weight measurement availability, and monthly clinical documentation across the high response group, moderate response group, late response group, weight regain group and the minimal weight-loss group.

| DRUG<br>(Active Compound) | BRAND<br>(Marketed Formulation) | Weight-Loss Outcome Category | Duration of prescriptions per patient in months, Mean (SD) | Duration of prescriptions per patient in months, Median [IQR] | Number of prescriptions per patient, Mean (SD) | Number of prescriptions per patient, Median [IQR] | Follow-up duration in days, Mean (SD) | Follow-up duration in days, Median [IQR] | Number of follow-up events per patient, Mean (SD) | Number of follow-up events per patient, Median [IQR] | Post-GLP1RA Weight Measurements, Mean (SD) | Post-GLP1RA Weight Measurements, Median [IQR] | Post-treatment monthly documents per patient, Mean (SD) | Post-treatment monthly documents per patient, Median [IQR] |
| --- | --- | --- | --- | --- | --- | --- | --- | --- | --- | --- | --- | --- | --- | --- |
| Semaglutide | (Ozempic/Wegovy/Rybelsus) | High Response Group | 12.04 (6.52) | 11.46 [7.11, 16.56] | 12.25 (8.53) | 10 [6, 16] | 501.46 (198.68) | 500 [354, 637] | 805.1 (1843.7) | 356 [201, 718] | 10.84 (12.09) | 8 [5, 13] | 6.0 (14.6) | 3 [2, 6] |
| Semaglutide | (Ozempic/Wegovy/Rybelsus) | Moderate Response Group | 9.45 (5.93) | 8.19 [4.99, 12.71] | 9.50 (6.35) | 8 [5, 12] | 409.44 (194.60) | 375 [266, 531] | 565.1 (1052.7) | 290 [163, 553] | 8.47 (7.31) | 6 [4, 10] | 5.7 (10.5) | 3 [2, 6] |
| Semaglutide | (Ozempic/Wegovy/Rybelsus) | Weight Regain Group | 11.31 (6.77) | 11.47 [5.61, 15.83] | 9.79 (6.99) | 8 [5, 12] | 570.41 (144.15) | 537 [457, 674] | 1233.7 (2499.9) | 499 [244, 989] | 14.00 (12.74) | 10 [7, 16] | 8.3 (20.0) | 4 [2, 8] |
| Semaglutide | (Ozempic/Wegovy/Rybelsus) | Late Response Group | 14.73 (6.71) | 15.38 [10.09, 19.84] | 10.80 (7.34) | 9 [6, 14] | 630.48 (127.45) | 629 [543, 724] | 801.0 (1688.2) | 376 [240, 693] | 11.13 (9.48) | 8 [6, 13] | 4.8 (7.1) | 3 [1, 6] |
| Semaglutide | (Ozempic/Wegovy/Rybelsus) | Minimal Weight-Loss Group | 12.81 (6.57) | 12.98 [7.92, 17.28] | 8.85 (6.23) | 8 [4, 12] | 585.78 (135.60) | 572 [474, 691] | 608.8 (1168.9) | 335 [212, 574] | 9.94 (7.46) | 8 [5, 12] | 5.2 (8.5) | 3 [2, 6] |
| Tirzepatide | (Mounjaro/Zepbound) | High Response Group | 12.90 (6.12) | 12.18 [8.60, 16.95] | 16.82 (11.16) | 15 [9, 22] | 491.06 (187.27) | 453 [352, 625] | 617.8 (1349.5) | 305 [181, 585] | 9.44 (9.19) | 7 [5, 11] | 4.9 (9.1) | 3 [2, 5] |
| Tirzepatide | (Mounjaro/Zepbound) | Moderate Response Group | 11.27 (6.09) | 10.29 [7.03, 14.62] | 13.51 (9.24) | 12 [7, 17] | 441.70 (183.43) | 399 [306, 560] | 504.4 (957.7) | 276 [164, 504] | 8.06 (7.10) | 6 [4, 9] | 5.0 (8.4) | 3 [2, 6] |
| Tirzepatide | (Mounjaro/Zepbound) | Weight Regain Group | 12.97 (7.07) | 13.13 [6.86, 18.14] | 13.62 (9.88) | 11 [6, 19] | 580.63 (147.98) | 552 [457, 693] | 969.2 (2254.7) | 468 [268, 928] | 13.16 (11.65) | 10 [7, 16] | 6.6 (13.6) | 4 [2, 7] |
| Tirzepatide | (Mounjaro/Zepbound) | Late Response Group | 15.90 (6.66) | 16.69 [12.54, 20.44] | 13.34 (9.52) | 12 [6, 18] | 631.41 (137.83) | 630 [528, 732] | 679.2 (1098.7) | 393 [242, 715] | 10.29 (8.05) | 8 [6, 12] | 4.2 (5.7) | 3 [1, 5] |
| Tirzepatide | (Mounjaro/Zepbound) | Minimal Weight-Loss Group | 13.94 (6.94) | 14.05 [9.13, 18.60] | 11.02 (8.63) | 8 [4, 15] | 578.62 (142.68) | 558 [463, 683] | 526.5 (978.5) | 314 [207, 530] | 8.84 (6.42) | 7 [5, 11] | 4.8 (7.6) | 3 [1, 6] |

**Table S6.**
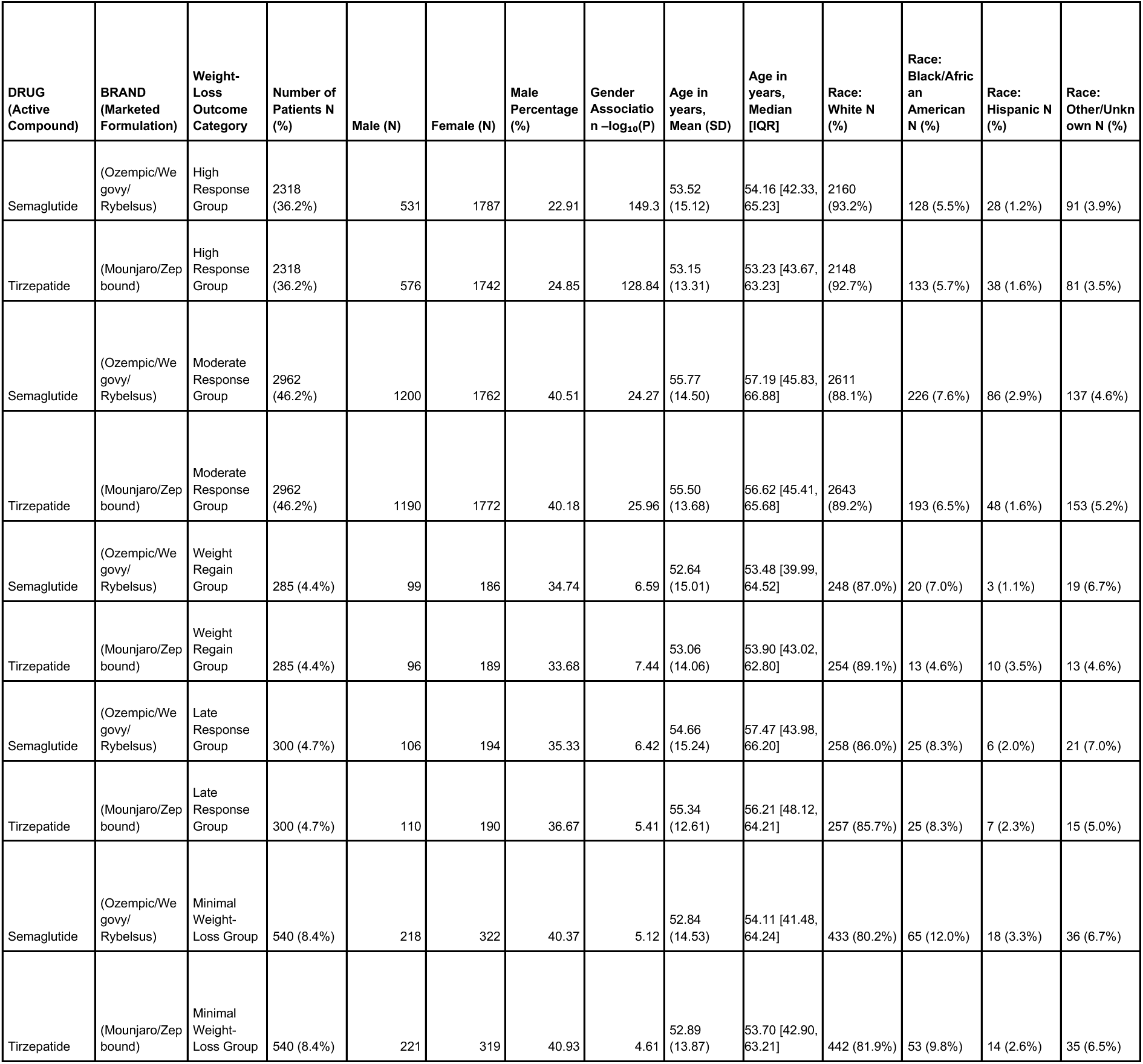
Demographic characteristics of response-group-specific propensity-matched tirzepatide vs semaglutide cohorts across the high response group, moderate response group, late response group, weight regain group and the minimal weight-loss group.

**Table S7.** Baseline clinical characteristics and data availability in response-group-specific propensity-matched tirzepatide versus semaglutide cohorts: Type 2 diabetes, BMI, weight distributions, weight measurement availability, and monthly clinical documentation across the high response group, moderate response group, late response group, weight regain group and the minimal weight-loss group.

| DRUG<br>(Active Compound) | BRAND<br>(Marketed Formulation) | Weight-Loss Outcome Category | With T2DM (N) | Without T2DM (N) | T2DM Percentage (%) | T2DM Association – log<sub>10</sub>(P) | Baseline BMI in kg/m<sup>2</sup>, Mean (SD) | Baseline BMI in kg/m<sup>2</sup>, Median [IQR] | Baseline Weight in kg, Mean (SD) | Baseline Weight in kg, Median [IQR] | Pre-GLP1RA Weight Measurements, Mean (SD) | Pre-GLP1RA Weight Measurements, Median [IQR] | Baseline monthly documentation per patient, Mean (SD) | Baseline monthly documentation per patient, Median [IQR] |
| --- | --- | --- | --- | --- | --- | --- | --- | --- | --- | --- | --- | --- | --- | --- |
| Semaglutide | (Ozempic /Wegovy/ Rybelsus) | High Response Group | 621 | 1697 | 26.79 | 109.91 | 36.93 (5.94) | 36.44 [32.56, 41.55] | 105.21 (22.31) | 101.79 [88.47, 118.73] | 38.18 (37.67) | 27 [13, 50] | 6.7 (15.4) | 3 [2, 6] |
| Tirzepatide | (Mounjaro/Zepbound) | High Response Group | 658 | 1660 | 28.39 | 95.47 | 37.52 (6.30) | 37.18 [32.82, 42.13] | 108.19 (23.13) | 104.96 [90.59, 122.66] | 34.29 (34.46) | 25 [13, 45] | 6.2 (14.6) | 3 [2, 6] |
| Semaglutide | (Ozempic /Wegovy/ Rybelsus) | Moderate Response Group | 1015 | 1947 | 34.27 | 65.01 | 36.78 (6.47) | 36.34 [32.14, 41.31] | 108.14 (23.76) | 105.37 [90.77, 123.11] | 37.87 (37.14) | 28 [12, 52] | 6.8 (15.2) | 3 [2, 7] |
| Tirzepatide | (Mounjaro/Zepbound) | Moderate Response Group | 1003 | 1959 | 33.86 | 68.35 | 36.93 (6.47) | 36.57 [32.23, 41.40] | 108.97 (24.09) | 106.49 [91.21, 123.34] | 34.15 (32.20) | 26 [12, 46] | 6.2 (13.5) | 3 [2, 6] |
| Semaglutide | (Ozempic /Wegovy/ Rybelsus) | Weight Regain Group | 85 | 200 | 29.82 | 11.02 | 35.60 (6.76) | 34.17 [30.90, 40.07] | 101.72 (21.30) | 99.86 [86.17, 112.62] | 36.44 (36.88) | 27 [13, 48] | 8.0 (19.0) | 3 [2, 8] |
| Tirzepatide | (Mounjaro/Zepbound) | Weight Regain Group | 88 | 197 | 30.88 | 9.97 | 35.39 (6.22) | 34.72 [31.07, 38.78] | 101.46 (22.04) | 97.83 [86.50, 114.88] | 34.37 (31.73) | 28 [13, 43] | 6.8 (13.3) | 3 [2, 7] |
| Semaglutide | (Ozempic /Wegovy/ Rybelsus) | Late Response Group | 104 | 196 | 34.67 | 6.96 | 37.91 (7.31) | 37.52 [33.02, 42.16] | 111.71 (24.09) | 108.87 [93.87, 126.39] | 32.18 (29.91) | 26 [13, 42] | 6.4 (13.5) | 3 [2, 6] |
| Tirzepatide | (Mounjaro/Zepbound) | Late Response Group | 107 | 193 | 35.67 | 6.16 | 37.61 (6.77) | 37.37 [32.93, 41.70] | 111.40 (24.88) | 107.39 [92.83, 126.81] | 32.16 (33.37) | 23 [12, 44] | 7.2 (16.1) | 3 [2, 7] |
| Semaglutide | (Ozempic /Wegovy/ Rybelsus) | Minimal Weight-Loss Group | 178 | 362 | 32.96 | 14.62 | 38.40 (7.99) | 37.93 [32.42, 43.40] | 113.66 (27.15) | 110.86 [93.15, 130.84] | 33.49 (31.66) | 24 [12, 48] | 7.3 (15.7) | 3 [2, 7] |
| Tirzepatide | (Mounjaro/Zepbound) | Minimal Weight-Loss Group | 168 | 372 | 31.11 | 17.78 | 37.97 (8.97) | 37.34 [31.48, 43.29] | 112.43 (29.37) | 109.36 [91.52, 132.14] | 29.53 (26.27) | 23 [10, 40] | 6.9 (14.0) | 3 [2, 7] |

**Table S8.** Post-treatment characteristics in response-group-specific propensity-matched tirzepatide versus semaglutide cohorts: prescription duration and frequency, follow-up duration, weight measurement availability, and monthly clinical documentation across the high response group, moderate response group, late response group, weight regain group and the minimal weight-loss group.

| DRUG<br>(Active Compound) | BRAND<br>(Marketed Formulation) | Weight-Loss Outcome Category | Duration of prescriptions per patient in months, Mean (SD) | Duration of prescriptions per patient in months, Median [IQR] | Number of prescriptions per patient, Mean (SD) | Number of prescriptions per patient, Median [IQR] | Follow-up duration in days, Mean (SD) | Follow-up duration in days, Median [IQR] | Number of follow-up events per patient, Mean (SD) | Number of follow-up events per patient, Median [IQR] | Post-GLP1RA Weight Measurements, Mean (SD) | Post-GLP1RA Weight Measurements, Median [IQR] | Post-treatment monthly documents per patient, Mean (SD) | Post-treatment monthly documents per patient, Median [IQR] |
| --- | --- | --- | --- | --- | --- | --- | --- | --- | --- | --- | --- | --- | --- | --- |
| Semaglutide | (Ozempic/Wegovy/Rybelsus) | High Response Group | 12.50 (6.55) | 12.00 [7.49, 17.10] | 12.67 (8.80) | 11 [6, 16] | 517.68 (187.59) | 517 [377, 646] | 961.3 (2249.5) | 415 [233, 860] | 12.23 (11.97) | 9 [6, 14] | 6.1 (14.1) | 3 [2, 6] |
| Tirzepatide | (Mounjaro/Zepbound) | High Response Group | 14.06 (6.02) | 13.51 [10.41, 18.04] | 17.41 (11.53) | 16 [10, 22] | 534.43 (178.91) | 516 [407, 647] | 741.3 (1564.0) | 376 [220, 718] | 10.88 (11.07) | 8 [5, 13] | 5.0 (8.7) | 3 [2, 6] |
| Semaglutide | (Ozempic/Wegovy/Rybelsus) | Moderate Response Group | 9.16 (5.84) | 7.80 [4.72, 12.57] | 9.61 (6.45) | 8 [5, 13] | 394.35 (189.87) | 356 [252, 515] | 619.4 (1235.8) | 299 [167, 609] | 9.08 (9.21) | 6 [5, 11] | 5.8 (10.7) | 3 [2, 6] |
| Tirzepatide | (Mounjaro/Zepbound) | Moderate Response Group | 10.16 (5.94) | 9.04 [5.93, 13.25] | 12.86 (8.73) | 11 [7, 17] | 397.74 (185.78) | 356 [260, 502] | 531.1 (1012.9) | 282 [164, 537] | 8.32 (7.77) | 6 [4, 10] | 5.2 (8.8) | 3 [2, 6] |
| Semaglutide | (Ozempic/Wegovy/Rybelsus) | Weight Regain Group | 12.23 (6.95) | 12.67 [6.83, 16.50] | 10.36 (7.62) | 8 [6, 14] | 591.73 (150.19) | 567 [470, 730] | 1104.8 (1869.9) | 562 [282, 1083] | 14.02 (10.30) | 11 [7, 17] | 7.6 (17.9) | 4 [2, 7] |
| Tirzepatide | (Mounjaro/Zepbound) | Weight Regain Group | 12.73 (7.34) | 12.71 [6.62, 17.89] | 13.53 (9.33) | 12 [6, 19] | 583.86 (151.71) | 550 [459, 708] | 1012.7 (2370.6) | 500 [293, 943] | 13.70 (11.89) | 10 [7, 16] | 6.5 (12.0) | 3 [2, 7] |
| Semaglutide | (Ozempic/Wegovy/Rybelsus) | Late Response Group | 14.07 (6.90) | 14.74 [8.70, 19.05] | 10.36 (7.26) | 8 [5, 13] | 628.18 (126.36) | 634 [538, 724] | 900.8 (1852.0) | 464 [299, 834] | 11.96 (9.74) | 9 [6, 15] | 5.0 (7.8) | 3 [2, 5] |
| Tirzepatide | (Mounjaro/Zepbound) | Late Response Group | 15.82 (6.67) | 16.19 [12.59, 20.49] | 13.15 (9.01) | 12 [6, 18] | 626.63 (137.65) | 618 [520, 730] | 803.1 (1124.5) | 472 [276, 842] | 11.49 (8.82) | 8 [6, 14] | 4.4 (6.1) | 3 [1, 5] |
| Semaglutide | (Ozempic/Wegovy/Rybelsus) | Minimal Weight-Loss Group | 12.34 (6.28) | 12.65 [7.76, 16.71] | 9.14 (6.51) | 8 [4, 11] | 573.75 (139.89) | 554 [456, 681] | 666.3 (1391.8) | 367 [232, 654] | 11.04 (8.17) | 8 [6, 13] | 5.2 (8.2) | 3 [2, 6] |
| Tirzepatide | (Mounjaro/Zepbound) | Minimal Weight-Loss Group | 13.70 (6.99) | 13.80 [8.97, 18.49] | 11.25 (9.18) | 8 [5, 15] | 571.72 (139.85) | 553 [463, 665] | 634.6 (1217.7) | 354 [225, 618] | 9.83 (7.30) | 8 [5, 12] | 5.2 (8.8) | 3 [2, 6] |

**Table S9:** Adverse events across propensity-matched high response groups of Tirzepatide vs Semaglutide (N=2,318)

| Adverse Event | Tirzepatide Prevalence (%) | Semaglutide Prevalence (%) | Tirzepatide Patients (N) | Semaglutide Patients (N) | Neg Log P-value |
| --- | --- | --- | --- | --- | --- |
| Nausea | 27.22 | 31.41 | 631 | 728 | 2.7094 |
| Fatigue | 26.1 | 29.72 | 605 | 689 | 2.1820 |
| Headache | 23.51 | 28.43 | 545 | 659 | 3.8133 |
| Constipation | 16.91 | 20.19 | 392 | 468 | 2.3372 |
| Diarrhea | 16.87 | 19.24 | 391 | 446 | 1.4065 |
| Vomiting | 12.81 | 17.13 | 297 | 397 | 4.3378 |
| Abdominal Pain | 14.45 | 16.52 | 335 | 383 | 1.2488 |
| Dizziness | 14.84 | 15.92 | 344 | 369 | 0.4834 |
| Heartburn | 4.23 | 4.06 | 98 | 94 | 0.0836 |
| Hypoglycemia | 3.67 | 3.67 | 85 | 85 | 0.0000 |
| Dyspepsia | 0.69 | 1.6 | 16 | 37 | 2.2421 |
| Decreased Appetite | 0.78 | 0.95 | 18 | 22 | 0.1981 |
| Flatulence | 0.73 | 0.86 | 17 | 20 | 0.1300 |
| Belching | 0.47 | 0.22 | 11 | 5 | 0.6767 |
| Abdominal Distension | 0.22 | 0.22 | 5 | 5 | 0.0000 |
| Gastroenteritis | 0.22 | 0.22 | 5 | 5 | 0.0000 |

**Table S10:**
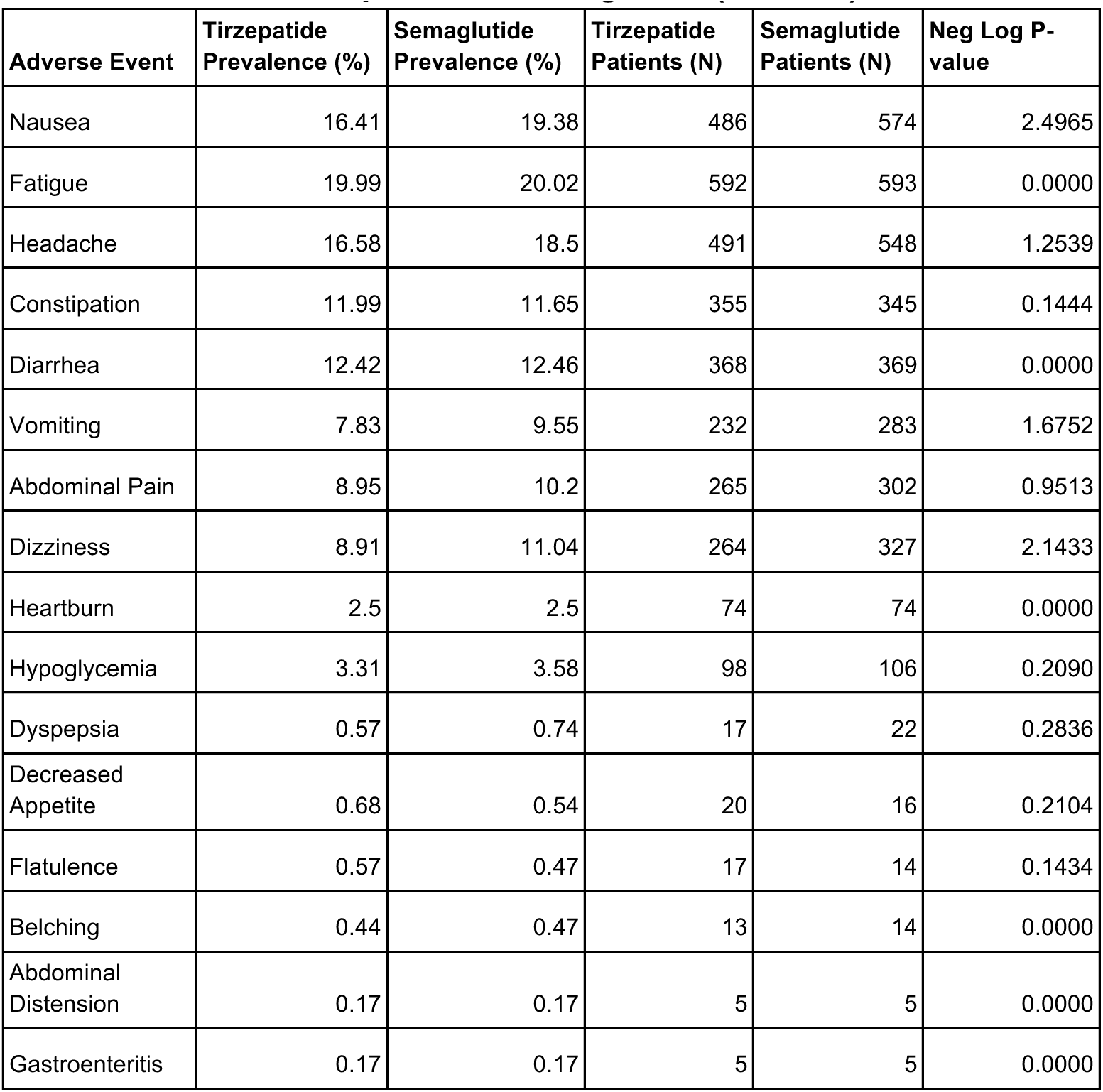
Adverse events across propensity-matched moderate response groups of Tirzepatide vs Semaglutide (N=2,962)

**Table S11:** Adverse events across propensity-matched minimal weight-loss groups of Tirzepatide vs Semaglutide (N=540)

| Adverse Event | Tirzepatide Prevalence (%) | Semaglutide Prevalence (%) | Tirzepatide Patients (N) | Semaglutide Patients (N) | Neg Log P-value |
| --- | --- | --- | --- | --- | --- |
| Nausea | 18.7 | 22.41 | 101 | 121 | 0.8167 |
| Fatigue | 22.96 | 23.89 | 124 | 129 | 0.1114 |
| Headache | 19.44 | 27.96 | 105 | 151 | 2.8920 |
| Constipation | 8.7 | 9.63 | 47 | 52 | 0.1719 |
| Diarrhea | 15 | 12.78 | 81 | 69 | 0.4774 |
| Vomiting | 9.26 | 10.56 | 50 | 57 | 0.2667 |
| Abdominal Pain | 11.67 | 10.37 | 63 | 56 | 0.2519 |
| Dizziness | 10.19 | 12.41 | 55 | 67 | 0.5371 |
| Heartburn | 0.93 | 1.85 | 5 | 10 | 0.5253 |
| Hypoglycemia | 0.93 | 0.93 | 5 | 5 | 0.0000 |
| Dyspepsia | 0.93 | 0.93 | 5 | 5 | 0.0000 |
| Decreased Appetite | 0.93 | 0.93 | 5 | 5 | 0.0000 |
| Flatulence | 0.93 | 0.93 | 5 | 5 | 0.0000 |
| Belching | 0.93 | 0.93 | 5 | 5 | 0.0000 |
| Abdominal Distension | 0 | 0 | 0 | 0 | 0.0000 |
| Gastroenteritis | 0.93 | 0.93 | 5 | 5 | 0.0000 |

**Table S12:**
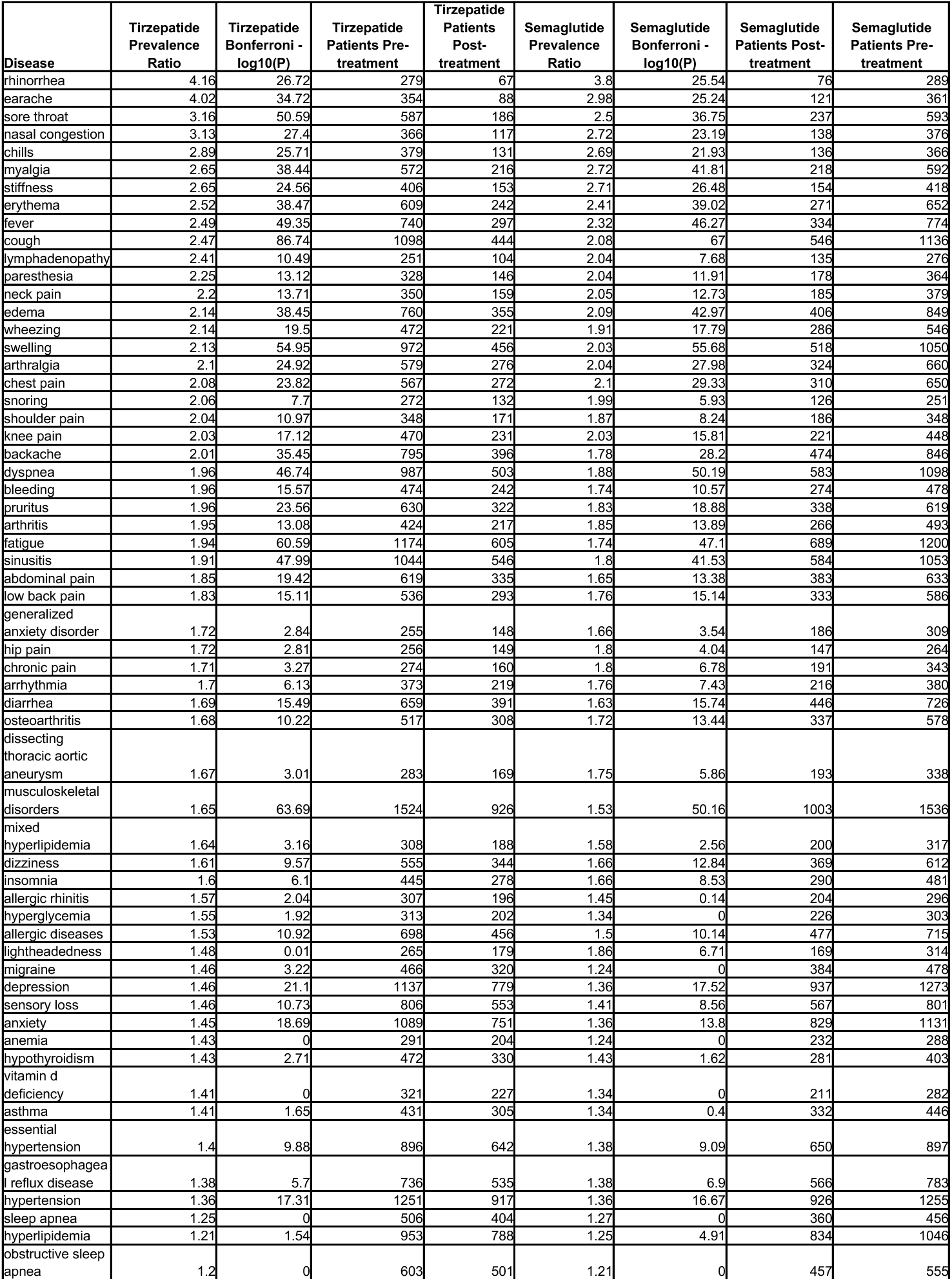
Pre- and post-treatment disease prevalence across propensity-matched high response groups of Tirzepatide vs Semaglutide (N=2,318)

**Table S13:**
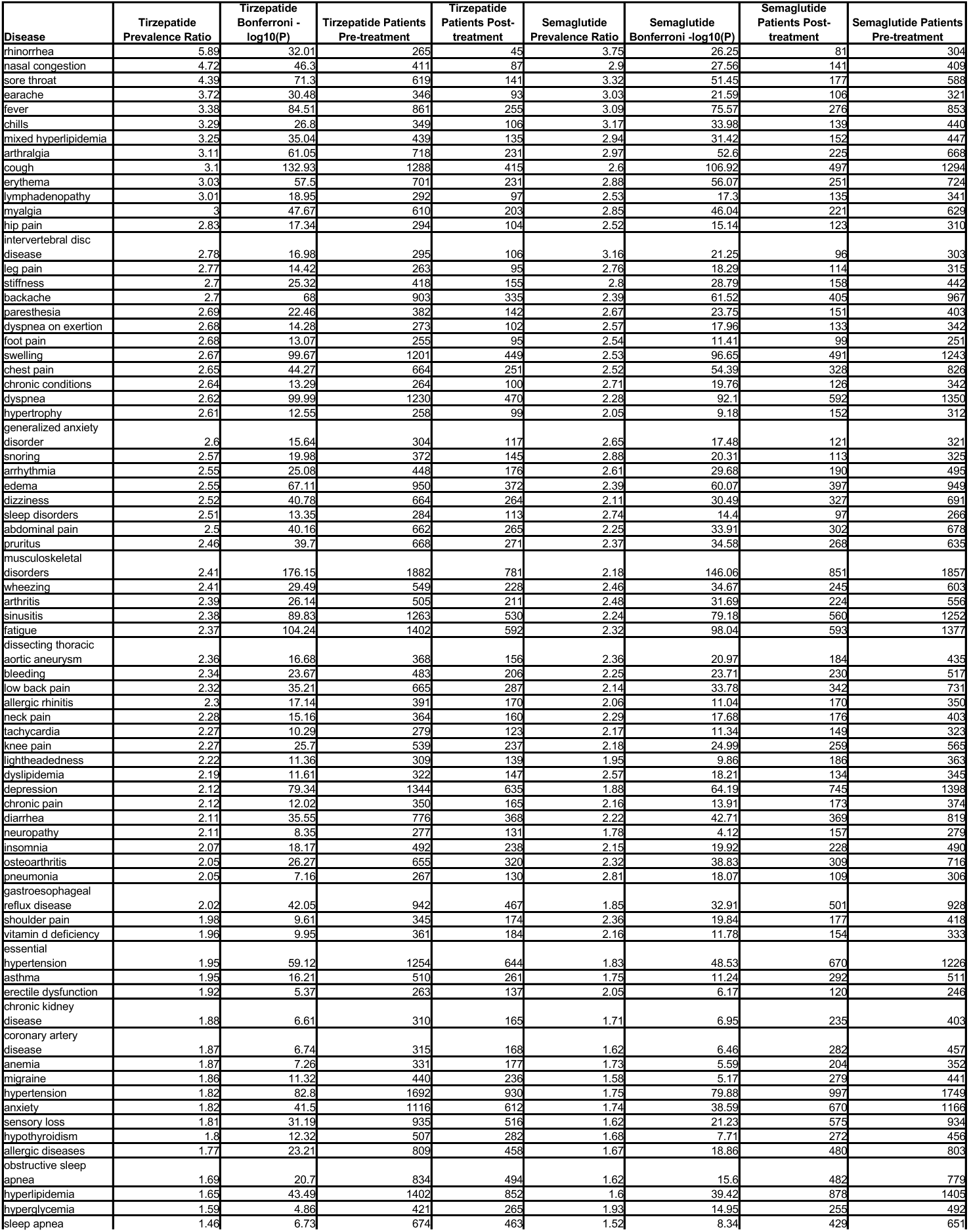
Pre- and post-treatment disease prevalence across propensity-matched moderate response groups of Tirzepatide vs Semaglutide (N=2,692)

**Table S14:**
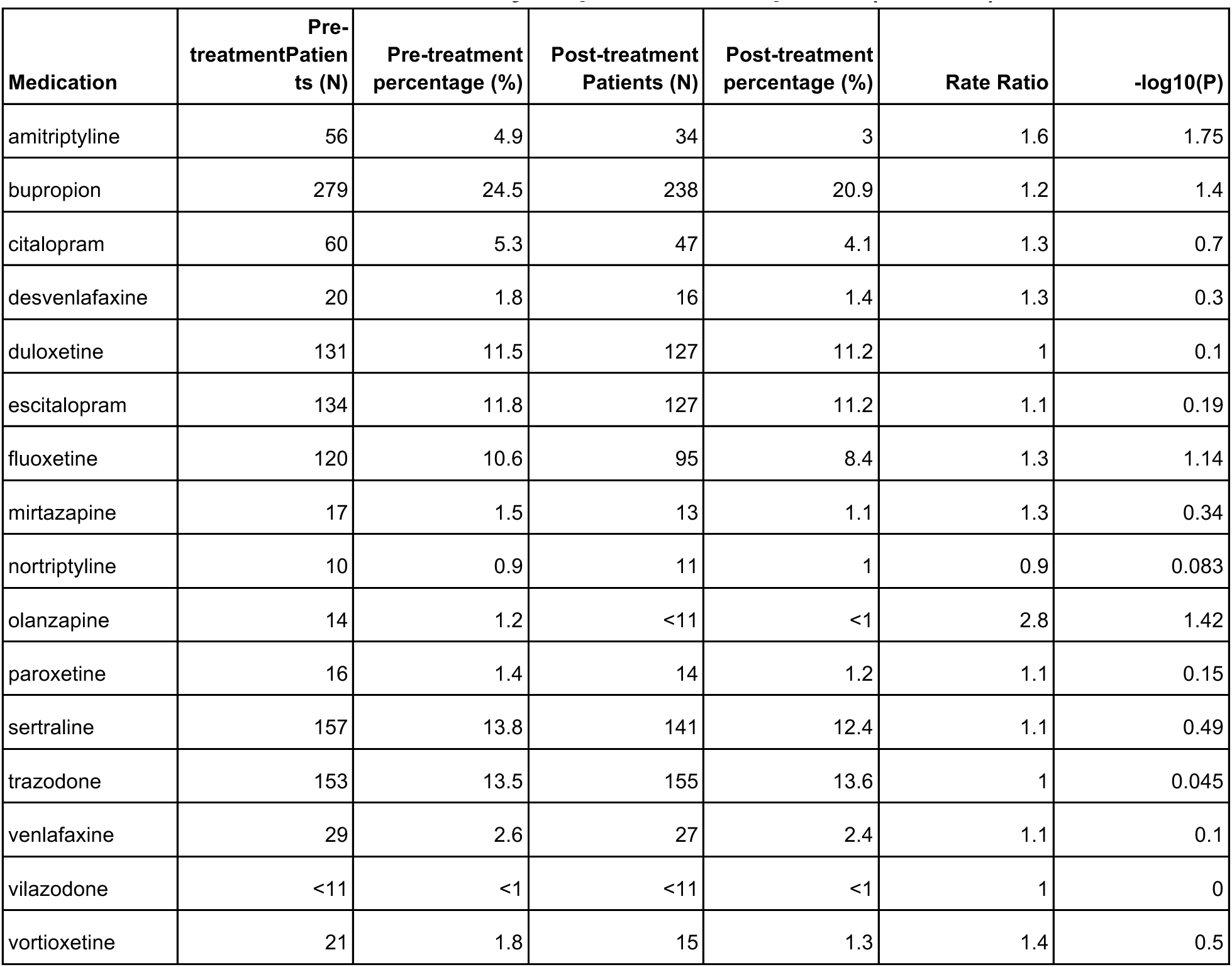
Two year pre- versus post-treatment medication prevalence for selected depression medications among the Tirzepatide high response group with depression mentions in the two year pre-treatment period (N=1,137)

| Medication | Pre-treatment Patients (N) | Pre-treatment percentage (%) | Post-treatment Patients (N) | Post-treatment percentage (%) | Rate Ratio | -log10(P) |
| --- | --- | --- | --- | --- | --- | --- |
| amitriptyline | 56 | 4.9 | 34 | 3 | 1.6 | 1.75 |
| bupropion | 279 | 24.5 | 238 | 20.9 | 1.2 | 1.4 |
| citalopram | 60 | 5.3 | 47 | 4.1 | 1.3 | 0.7 |
| desvenlafaxine | 20 | 1.8 | 16 | 1.4 | 1.3 | 0.3 |
| duloxetine | 131 | 11.5 | 127 | 11.2 | 1 | 0.1 |
| escitalopram | 134 | 11.8 | 127 | 11.2 | 1.1 | 0.19 |
| fluoxetine | 120 | 10.6 | 95 | 8.4 | 1.3 | 1.14 |
| mirtazapine | 17 | 1.5 | 13 | 1.1 | 1.3 | 0.34 |
| nortriptyline | 10 | 0.9 | 11 | 1 | 0.9 | 0.083 |
| olanzapine | 14 | 1.2 | <11 | <1 | 2.8 | 1.42 |
| paroxetine | 16 | 1.4 | 14 | 1.2 | 1.1 | 0.15 |
| sertraline | 157 | 13.8 | 141 | 12.4 | 1.1 | 0.49 |
| trazodone | 153 | 13.5 | 155 | 13.6 | 1 | 0.045 |
| venlafaxine | 29 | 2.6 | 27 | 2.4 | 1.1 | 0.1 |
| vilazodone | <11 | <1 | <11 | <1 | 1 | 0 |
| vortioxetine | 21 | 1.8 | 15 | 1.3 | 1.4 | 0.5 |

**Table S15:**
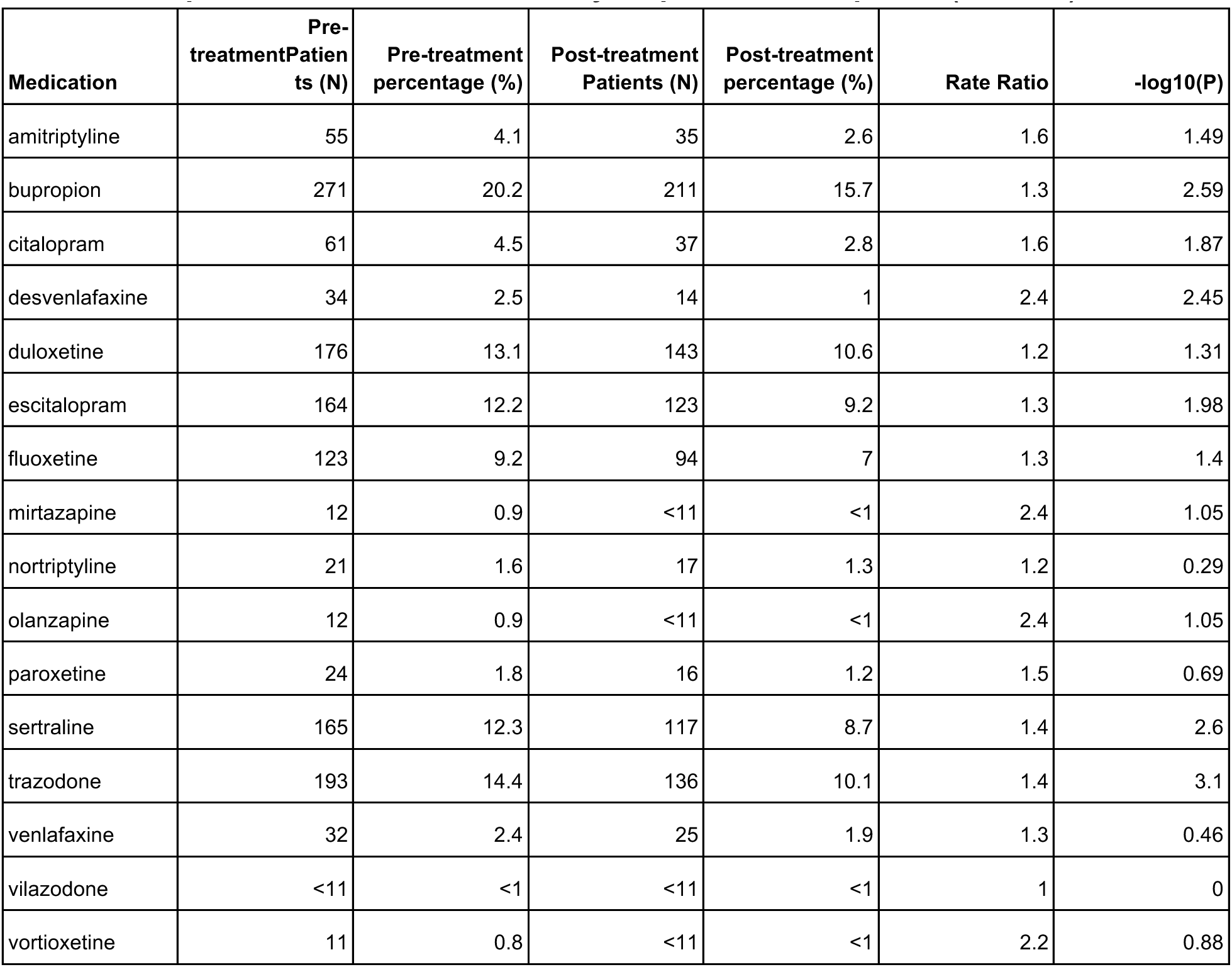
Two year pre- versus post-treatment medication prevalence for selected depression medications among the Tirzepatide moderate response group with depression mentions in the two year pre-treatment period (N=1,344)

| Medication | Pre-treatment Patients (N) | Pre-treatment percentage (%) | Post-treatment Patients (N) | Post-treatment percentage (%) | Rate Ratio | -log10(P) |
| --- | --- | --- | --- | --- | --- | --- |
| amitriptyline | 55 | 4.1 | 35 | 2.6 | 1.6 | 1.49 |
| bupropion | 271 | 20.2 | 211 | 15.7 | 1.3 | 2.59 |
| citalopram | 61 | 4.5 | 37 | 2.8 | 1.6 | 1.87 |
| desvenlafaxine | 34 | 2.5 | 14 | 1 | 2.4 | 2.45 |
| duloxetine | 176 | 13.1 | 143 | 10.6 | 1.2 | 1.31 |
| escitalopram | 164 | 12.2 | 123 | 9.2 | 1.3 | 1.98 |
| fluoxetine | 123 | 9.2 | 94 | 7 | 1.3 | 1.4 |
| mirtazapine | 12 | 0.9 | <11 | <1 | 2.4 | 1.05 |
| nortriptyline | 21 | 1.6 | 17 | 1.3 | 1.2 | 0.29 |
| olanzapine | 12 | 0.9 | <11 | <1 | 2.4 | 1.05 |
| paroxetine | 24 | 1.8 | 16 | 1.2 | 1.5 | 0.69 |
| sertraline | 165 | 12.3 | 117 | 8.7 | 1.4 | 2.6 |
| trazodone | 193 | 14.4 | 136 | 10.1 | 1.4 | 3.1 |
| venlafaxine | 32 | 2.4 | 25 | 1.9 | 1.3 | 0.46 |
| vilazodone | <11 | <1 | <11 | <1 | 1 | 0 |
| vortioxetine | 11 | 0.8 | <11 | <1 | 2.2 | 0.88 |

**Table S16:**
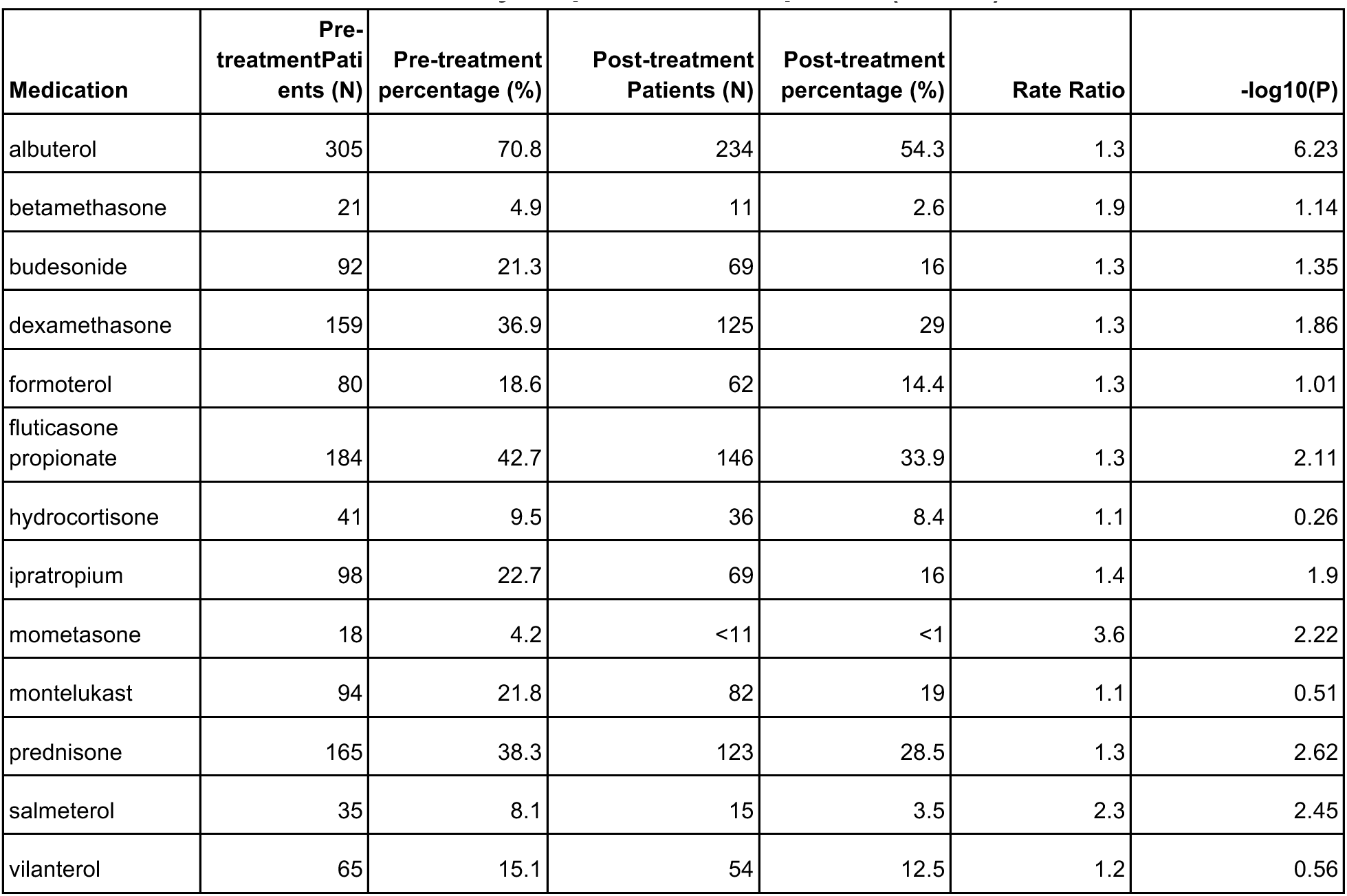
Two year pre- versus post-treatment medication prevalence for selected asthma medications among the Tirzepatide high response group with asthma mentions in the two year pre-treatment period (N=431)

| Medication | Pre-treatment Patients (N) | Pre-treatment percentage (%) | Post-treatment Patients (N) | Post-treatment percentage (%) | Rate Ratio | -log10(P) |
| --- | --- | --- | --- | --- | --- | --- |
| albuterol | 305 | 70.8 | 234 | 54.3 | 1.3 | 6.23 |
| betamethasone | 21 | 4.9 | 11 | 2.6 | 1.9 | 1.14 |
| budesonide | 92 | 21.3 | 69 | 16 | 1.3 | 1.35 |
| dexamethasone | 159 | 36.9 | 125 | 29 | 1.3 | 1.86 |
| formoterol | 80 | 18.6 | 62 | 14.4 | 1.3 | 1.01 |
| fluticasone propionate | 184 | 42.7 | 146 | 33.9 | 1.3 | 2.11 |
| hydrocortisone | 41 | 9.5 | 36 | 8.4 | 1.1 | 0.26 |
| ipratropium | 98 | 22.7 | 69 | 16 | 1.4 | 1.9 |
| mometasone | 18 | 4.2 | <11 | <1 | 3.6 | 2.22 |
| montelukast | 94 | 21.8 | 82 | 19 | 1.1 | 0.51 |
| prednisone | 165 | 38.3 | 123 | 28.5 | 1.3 | 2.62 |
| salmeterol | 35 | 8.1 | 15 | 3.5 | 2.3 | 2.45 |
| vilanterol | 65 | 15.1 | 54 | 12.5 | 1.2 | 0.56 |

**Table S17:** Two year pre- versus post-treatment medication prevalence for selected asthma medications among the Tirzepatide moderate response group with asthma mentions in the two year pre-treatment period (N=510)

| **Medication** | **Pre-treatment Patients (N)** | **Pre-treatment percentage (%)** | **Post-treatment Patients (N)** | **Post-treatment percentage (%)** | **Rate Ratio** | **-log10(P)** |
| --- | --- | --- | --- | --- | --- | --- |
| albuterol | 350 | 68.6 | 217 | 42.5 | 1.6 | 16.28 |
| betamethasone | 21 | 4.1 | 13 | 2.5 | 1.6 | 0.79 |
| budesonide | 92 | 21.3 | 69 | 16 | 1.3 | 1.35 |
| dexamethasone | 191 | 37.5 | 139 | 27.3 | 1.4 | 3.3 |
| formoterol | 76 | 14.9 | 53 | 10.4 | 1.4 | 1.52 |
| fluticasone propionate | 220 | 43.1 | 144 | 28.2 | 1.5 | 6.17 |
| hydrocortisone | 49 | 9.6 | 29 | 5.7 | 1.7 | 1.73 |
| ipratropium | 98 | 19.2 | 73 | 14.3 | 1.3 | 1.44 |
| mometasone | 25 | 4.9 | <11 | <1 | 5 | 3.68 |
| montelukast | 128 | 25.1 | 96 | 18.8 | 1.3 | 1.81 |
| prednisone | 199 | 39 | 110 | 21.6 | 1.8 | 8.88 |
| salmeterol | 43 | 8.4 | 23 | 4.5 | 1.9 | 1.96 |
| vilanterol | 64 | 12.5 | 48 | 9.4 | 1.3 | 0.96 |

